# Optimal sample pooling: an efficient tool against SARS-CoV-2

**DOI:** 10.1101/2020.07.03.20145953

**Authors:** Saurabh Goyal, Priti Bist, Rakesh Sharma

## Abstract

The SARS-CoV-2 pandemic situation has presented multiple imminent challenges to the nations around the globe. While health agencies around the world are exploring various options to contain the spread of this fatal viral infection, multiple strategies and guidelines are being issued to boost the fight against the disease. Identifying and isolating infected individuals at an early phase of the disease has been a very successful approach to stop the chain of transmission. But this approach faces a practical challenge of limited resources. Sample pooling solves this enigma by significantly improving the testing capacity and result turn around time while using no extra resources. However, the general sample pooling method also has the scope of significant improvements. This article describes a process to further optimize the resources with optimal sample pooling. This is a user-friendly technique, scalable on a national or international scale. A mathematical model has been built and validated for its performance using clinical data.

## Introduction

The rapid rise in SARS-CoV-2 positive cases poses a significant health threat to the world. In India, the case count is surging as Government has announced significant relaxations in curbs for Unlock 1.^1^

Identifying and isolating the positive cases in the very early phase has been a major successful approach to stop the spread of the disease.^2, 3^ To identify such cases, the World Health Organization has stressed on multiple occasions the significant role of sample testing.^4, 5^ Yet unfortunately, in the testing strategies of majority of the nations, either only symptomatic cases are included or are heavily favoured.^6^ With such approach, the patient count tracker will always find itself trailing the steps of the viral spread. Multiple scientific studies state that the disease can lay asymptomatic for the first 2 to 3 weeks, immediately followed by the period of heavy viral shedding just as symptoms onset.^7-13^ Thus, testing all of the suspected individuals will provide an ideal solution. However, a major challenge in implementing such approach is resource limitation.

Many strategies have been identified to release the constraint of resource limitation. One of such tried and tested approach is sample pooling.^14-19^ Indian Council of Medical Research (ICMR) and other world health agencies have also suggested sample pooling as a major strategy to increase the scope of the test.^20, 21^ This strategy helps in multiple ways, it reduces both cost and result time.

Sample pooling can be achieved primarily in two ways: Repeated pooling and one-time pooling.^19^ In repeated sample pooling, a sample pool which has tested positive is further broken down into sub-pools and the testing process is repeated till each of the individual samples with positive SARS-CoV-2 infection is identified. This method turns out to be more time and cost efficient as it can be designed by using algorithms like binary search tree.^19^ However, this method has serious practical limitations as – a) it is a complex procedure and requires training to implement and b) it shows significantly higher efficiency when the initial pool size is high. In contrast, during one-time pooling, only one level of pooling is done. If a pool tests positive, in the next step, each individual sample of the pool is tested for the infection. Hence, one-time pooling is a more practical approach than the repeated pooling.

While the current guidelines from ICMR state that up to 5 samples can be pooled,^20^ multiple studies have confirmed that the pooling size of up to 8 does not harm the specificity and the sensitivity of the test.^16^ The latest guideline has left it up to the lab to decide the optimal pool size. Multiple tools have been reported in the scientific community to find the optimal pool size.^15^ While most of such tools consider multiple parameters to arrive at the optimal pool size, complicated procedures avert the practical use.

The current paper proposes a practical approach to find the optimal pool size. This approach is simple, easy to implement on a national scale and is adaptable to the stage of the viral spread.

## Method

Testing facilities are either already designated to test samples from a specific geography or can be easily mapped. This correlation ensures that the prevalence rate of the lab remains a continuous function. Sample data verifies this hypothesis. The temporal variance in the prevalence rate is significantly lower for a testing facility level than at a country or even at a state level where the variance is extremely high.^23^ Hence, in order to determine the current prevalence rate and to forecast the same, the metrics should be calculated for individual testing facilities. The determination of sample pool size for each lab using its prevalence rate would yield desired efficiency and be easy to implement.

Following mathematical model describes the gain factor of the sample pooling against individual testing of each sample. Involved parameters are,

**Current prevalence rate**, the probability of a sample being positive is forecasted using the actual prevalence rates of the last one week, ***p***

Samples to be tested, ***n***

Sample pool size, ***x*** (min – 2, max – 8)

No of tests without sample pooling, **N**_**normal**_ **= n**,

No. of tests after sample pooling,

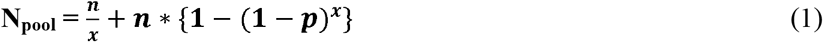

Gain factor,

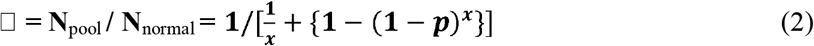

Using equation (2), gain factor values for different sample pool size and varying prevalence rate are simulated (Table 1).

**Table 1.** Gain factor simulation results for different sample pool size and varying prevalence rates

| GAIN FACTOR |  | Pool Size |  |  |  |  |  |  |
| --- | --- | --- | --- | --- | --- | --- | --- | --- |
|  |  | 2 | 3 | 4 | 5 | 6 | 7 | 8 |
| Prevalence Rate | 1% | 1.9 | 2.8 | 3.5 | 4.0 | 4.4 | 4.7 | 4.9 |
|  | 3% | 1.8 | 2.4 | 2.7 | 2.9 | 3.0 | 3.0 | 2.9 |
|  | 5% | 1.7 | 2.1 | 2.3 | 2.3 | 2.3 | 2.2 | 2.2 |
|  | 10% | 1.4 | 1.7 | 1.7 | 1.6 | 1.6 | 1.5 | 1.4 |
|  | 20% | 1.2 | 1.2 | 1.2 | 1.1 | 1.1 | 1.1 | 1.0 |

**Table 2.** Decision matrix for optimal pool size selection

| Min | Max | Result | Pool size |
| --- | --- | --- | --- |
| 0% | 1% | 4.0 | 5 |
| 1% | 3% | 2.9 | 5 |
| 3% | 5% | 2.3 | 5 |
| 5% | 10% | 1.7 | 4 |
| 10% | 20% | 1.2 | 4 |
| 20% | 100% | Do not Pool | Do not Pool |

In the situation of prevalence rate of 1% or lower, though higher sample pool size yields better gain factors, yet the risk of sample dilution outweighs the miniscule gains. Thus, the sample pool size of 5 is suitable during lower prevalence rates, less than 5%, while for higher prevalence rates, more than 5%, pool size of 4 provides the best efficiency. However, for prevalence rates above 20%, sample pooling should not be used.

## Discussion and future work

Strategizing to have a common sample pool size across the nation would not yield optimum results as the variance of prevalence rates is extremely high. Hence, sample pool size should be decided individually for a testing facility using the prevalence rates recorded by the same lab. Our results show that a period of 7 days can be used to forecast the prevalence rate of the next day. This prevalence rate can then be looked up on the decision matrix table to arrive at the optimal sample pool size. However, sample from the following categories should be excluded from the process:

1. A pool or an individual sample which has already been tested positive
2. Special testing scenarios, such as, retesting or testing of a specific group which is bound to be having much higher risk than the normal conditions.

Though the mathematical model has been tested with the clinical data, a clinical study to test the performance is being planned.

## Data Availability

The data used for the study is available in the public domain.

http://www.uhsr.ac.in/detail.aspx?artid=67&menuid=89.

## Supplementary Information

Prevalence rate variation of a lab in clinical setting.

## Notes

### Competing Interest Statement

The authors have declared no competing interest.

### Funding Statement

No funding was received.

### Author Declarations

Our research does not fall into the purview of any oversight body.

